# The causal association between maternal depression, anxiety and infection in pregnancy and neurodevelopmental disorders among 410,461 children- a population study using quasi-negative control cohorts and sibling analysis

**DOI:** 10.1101/2023.05.16.23290039

**Authors:** Holly Hope, Matthias Pierce, Hend Gabr, Maja R Radojčić, Eleanor Swift, Vicky P Taxiarchi, Kathryn M Abel

## Abstract

**Background:** To address if the long-standing association between maternal infection, depression/anxiety in pregnancy and offspring neurodevelopmental disorder (NDD) is causal we conducted two negative-control studies.

**Methods:** Four primary care cohorts of UK children (pregnancy, 1 and 2 years prior to pregnancy and siblings) born between 1st January 1990 to 31st December 2017 were constructed.

NDD included autism/autism spectrum disorder, attention deficit/hyperactivity disorder, intellectual disability, cerebral palsy, and epilepsy. Maternal exposures included depression/anxiety and/or infection. Maternal (age, smoking status, comorbidities, BMI (Body Mass Index), NDD); child (gender, ethnicity, birth year); and area-level (region and level of deprivation) confounders were captured.

The NDD incidence rate among 1) children exposed during or outside of pregnancy and 2) siblings discordant for exposure in pregnancy was compared using Cox-regression models, unadjusted and adjusted for confounders.

**Results:** The analysis included 410,461 children of 297,426 mothers and 2,793,018 person-years of follow-up with 8900 NDD cases (incidence rate=3.2 per 1000 person years). After adjustments, depression and anxiety consistently associated with NDD (pregnancy adjusted HR=1.58, 95%CI 1.46-1.72; 1-year adj.HR=1.49, 95%CI 1.39-1.60; 2-year adj.HR=1.62, 95%CI 1.50-1.74); and to a lesser extent, of infection (pregnancy adj. HR=1.16, 95%CI 1.10-1.22; 1-year adj.HR=1.20, 95%CI 1.14-1.27; 2-year adj.HR=1.19 95%CI 1.12-1.25). NDD risk did not differ among siblings discordant for pregnancy exposure to mental illness HR=0.97, 95%CI 0.77-1.21 or infection HR=0.99, 95%CI 0.90-1.08.

**Conclusions:** Triangulation of results from two negative control studies provided no evidence of a specific, and therefore causal, link between in-utero exposure to infection, common mental illness, and later development of NDD.

## Introduction

Maternal mental illness is an increasingly common phenomenon: up to 1 in 6 children is exposed during pregnancy (Abel et al., 2019a). Other pregnancy-related exposures, including small-for-gestational-age and prematurity are well-recognised antecedents of neurodevelopmental risk (Allotey et al., 2018; Arcangeli, Thilaganathan, Hooper, Khan, & Bhide, 2012) triggering postnatal infant monitoring and additional supports to families. This is not the case for infants exposed to maternal mental illness during pregnancy (Spittle & Treyvaud, 2016), despite the well-documented association between parental mental illness and offspring neurodevelopmental disorder (Ayano, Maravilla, & Alati, 2019; Chen et al., 2020). This may be because there remain important gaps in our knowledge about the mechanisms through which parental mental illness transmits neurodevelopmental risk (Ji-Xu & Vincent, 2020). Compared to paternal exposure, antenatal maternal mental illness has consistently stronger effects (Ayano et al., 2019), suggesting exposure during pregnancy might be important.

The ‘maternal immune activation’ or ‘MiA’ hypothesis is a popular mechanistic explanation for pregnancy-related risk transmission, whereby placental function is altered by maternal inflammatory humoral and cytokine responses causing atypical fetal development and neurodevelopmental disorder (Brown & Meyer, 2018). Most evidence for MiA derives from animal studies: inducing inflammatory responses in pregnant mice results in behavioural changes among offspring analogous to human autism spectrum disorder (ASD) (Weber-Stadlbauer et al., 2017). Candidate risks for MiA in humans include exposure to maternal stress/depression, maternal infection, or teratogenic drugs during pregnancy (Han et al., 2021). One case-control study undertook mid-gestational serum profiling of mothers of children with NDDs and suggested that chemokine and cytokine regulation were indeed altered in pregnancy in women of children with autism spectrum disorder (ASD) compared to controls; although case control studies are susceptible to selection and recall bias, and unmeasured confounding (Jones et al., 2017). Such studies do not consider whether maternal inflammatory regulation might be altered prior to pregnancy. To supplement animal and genetic studies, we can use large datasets, where it is unfeasible or unethical to randomise women to exposures, which can provide vital information about the transmission of neuropsychiatric risk(Hagberg, Robijn, & Jick, 2018), in particular the specific role of the maternal in-utero environment in the aetiology of NDD(Ji-Xu & Vincent, 2020; O’Connor & Ciesla, 2022; Rai et al., 2017). In a recent Swedish register study maternal hospitalisation for infection in pregnancy independently associated with risk of ASD, particularly ASD with intellectual disability (Lee et al., 2015); however, sibling analysis suggested significant unmeasured confounding (Brynge et al., 2022). Separately, in a Danish study similar small increased risk of NDD associated with maternal and paternal infection in pregnancy, which undermine MiA assumptions (Lydholm et al., 2019). In this analysis we triangulate two different negative control studies to assess causal inference of the MiA hypothesis in a large, whole UK population cohort and ask whether:

1. excess offspring neurodevelopment is followed by exposure to maternal mental illness or maternal infection during pregnancy compared to offspring exposed 1-2 years prior to pregnancy.
2. infants exposed to maternal mental illness or infection during pregnancy were at increased risk of NDDs compared to unexposed siblings.

## Methods

### Data source

We used CPRD GOLD which contains diagnosis, symptom, and prescription information from ∼500 UK general practices, representing ∼11 million patients (Herrett et al., 2015). In the UK, >98% of the population is registered with a primary care practice, which is free at the point of care and is a gateway to specialist medical services. Most common illnesses including depression and anxiety are diagnosed and managed entirely within primary care. The quality outcomes framework (QOF) includes new diagnosis of depression as an incentivised performance indicator for primary care physicians, making the CPRD a comprehensive dataset of the mental health status of registered patients (Khan, Harrison, & Rose, 2010; Department of Health and Social Care, 2022), suitable for capturing incidence and prevalence trends (Khan et al., 2010; Rait et al., 2009; Slee, Nazareth, Freemantle, & Horsfall, 2021). Validated linkages connect women to their pregnancies (Pregnancy Register (Minassian et al., 2019) and their children (mother-baby-link (Gallagher, Kousoulis, Williams, Valentine, & Myles, 2021). Three-quarters of English practices in CPRD also have linkage with hospital episodes data (HES) which has ICD10 diagnoses from clinical specialist visits and hospital admissions.

### Cohorts

The cohort consisted of children born between 01.01.90-31.12.17 registered at general practices that linked to secondary care datasets (Hospital Episodes (HES) in England (N= 715,287) and provided data of sufficient quality (N= 546,305). To create exposure and control cohorts, mothers could not transfer out of practice before the child’s date of registration at the practice (544,519). Children had to be linked to a single birth episode in the mother’s maternity record.

Three cohorts were constructed, designed to capture discrete periods of exposure to maternal mental illness or infection: during pregnancy, 1 year before pregnancy and between 1-2 years before pregnancy. Mothers in each cohort were included if they were registered at a practice for given lengths of time: 1) pregnancy cohort: registered for minimum of 280 days before the child’s date of birth; 2) 1-year before cohort: registered for minimum of 1 year and 280 days before the child’s date of birth; and 3) 2-years before cohort: registered for minimum of 2 years and 280 days before the child’s date of birth). After exclusions (eFig1), 410,461 children linked to 297,803 mothers were included in the pregnancy cohort: 352,993 children linked to 256.617 mothers in the 1 year before pregnancy cohort; and 303,016 children linked to 220,895 mothers in the 2 years before pregnancy cohort (eTable1). Median age of offspring at end of follow-up was 5 [IQR (interquartile range) 2 to 10].

### Outcomes

NDD included disorders which would be recorded in primary care in sufficient number from birth onwards with well understood neurodevelopmental consequences; attention deficit disorder (ADHD)/attention deficit disorder (ADD), autism/autism spectrum disorder, intellectual disability, cerebral palsy, and epilepsy. Cases were drawn from the following sources: primary care, outpatients, and hospital admissions. In addition, ADHD and ADD cases were identified from prescriptions data; dates from each source were captured and the earliest date used as the outcome event date (see etable2 for how NDD was coded, see <u>github</u> for codes).

### Exposures

Incident maternal depression and anxiety (using our algorithm which captures all depression and anxiety events in the mother’s primary care record (1) see (see eTable2 for algorithm and <u>githhub</u> for codes).

Infections – all records of viral or non-viral infections recorded in primary care see (see <u>github</u> for categories and codes).

Pregnancy duration was determined for each child in three ways: child’s date of birth minus

1) pregnancy start date in pregnancy register (n=344,500); 2) gestational age of child (n=26,244); where there was no recorded start date or gestational age; 3) 280 days before delivery date recorded in primary care record (n=41,380).

During each exposure period, exposure (yes/no) to each exposure variable was recorded. The trimester in which it occurred was also recorded. Prior to pregnancy, exposure (yes/no) to each variable was recorded where a mental illness/infection event occurred and 1 and 2 years prior to pregnancy start date.

### Measured confounders

During each exposure period, the following covariates were identified from primary care data: smoking status (current/ex-smoker/non-smoker); BMI, from height and weight recorded in and converted into categories (<18.5, <25, <30, <35, <40 and 40≥); alcohol or substance misuse (yes/no); and Charlson comorbidity index (CCI) (Charlson, Szatrowski, Peterson, & Gold, 1994) -a validated measure of health burden based on number and type of long-term health conditions, where higher scores indicate greater burden. If there were multiple different BMI or CCI scores recorded on the same date the mean score was used. Where there were multiple events within the exposure period the most severe score or category of confounder was selected. Other confounders that did not vary over time included: region of England, index of multiple deprivation, maternal history of NDD (yes/no) and birth order. Gender and ethnicity (Asian British/Black British/Mixed/Other/White British/Unknown) of child were extracted from the maternal record in HES and the child record in CPRD. Missing was treated as a separate category for categorical covariates.

Antidepressants and antibiotics were identified from the prescription’s dataset using the British National Formulary code, antidepressants include all prescriptions from chapter 4 section 3, and antibiotics from chapter 5 section 1. In pregnancy, exposure (yes/no) to each drug was recorded where it was prescribed in the 280 days prior to the child’s date of birth.

### Analysis

Each child was followed from the latest of these dates: their date of birth, date they were registered at their clinical practice. Follow-up ended at the earliest date of; the event of interest, the child’s date of death, 18^th^ birthday, the date they left their clinical practice, or the date their clinical practice stopped sharing data. Cox regression models were applied to the time-to-event data and hazard ratios measured the association between common mental illness or infection and each NDD outcome (any NDD, ASD, ADHD, intellectual disability, epilepsy, and cerebral palsy). Models were fitted on each cohort – pregnancy, 1 year before pregnancy and 2 years before pregnancy – adjusted for the same set of potential confounding variables. We adjusted the maternal infection analysis for maternal depression and the maternal depression analysis for maternal infection. Analyses were repeated for female and male offspring; and for trimester of exposure to interrogate gender and/or trimester effects. In the pregnancy cohort we adjusted the maternal infection analysis for maternal antibiotic use and the maternal depression analysis for maternal antidepressant use in pregnancy.

Finally, to account for unmeasured environmental and genetic confounding, we compared incidence rate of NDD, ADHD and ASD among siblings discordant for exposure to maternal common mental illness or infection. Discordant pairs were identified using the mother-baby link; where there was more than one possible discordant pair within a family, a pair was sampled at random. Adjusted estimates only included time-dependent variables (maternal age, smoking status, BMI, alcohol, or substance misuse, Charlson comorbidity index, gender, ethnicity, birth year and birth order of the child).

### Sensitivity analyses

We performed three sensitivity analyses. Potentially, NDD is misdiagnosed in primary care; therefore, we repeated analyses only using cases of NDD diagnosed in secondary care. Children born pre-term, who experience pregnancy and/or obstetric complications, are likely to be at increased risk of NDD; and may spend several weeks or months in hospital postnatally, delaying registration in primary care. However, late registration may also indicate family dysfunction or looked-after status of the child and inflate any effect of maternal mental illness or infection on risk of NDD. Therefore, we conducted a sensitivity analysis restricted to children registered at their general practice within 30 days of their date of birth (n=254,338).

It can be difficult to identify the end of mental illness episodes. Therefore, we repeated the main and sibling analyses by considering a woman’s incident mental illness to be episodic and women as exposed for 2 years after the incident event.

The association between maternal mental illness or infection and NDD also might be confounded by medication exposure. Therefore, we adjusted associations between mental illness, pregnancy and NDD for antidepressant use in pregnancy; risk estimates associated with infection were adjusted for antibiotic use in pregnancy.

## Results

### Characteristics of cohorts

The analysis included 410,461 children of 297,426 mothers and 2,793,018 person-years of follow-up with 8900 NDD cases (incidence rate = 3.2 per 1000 person years). Table 1 shows the rates of exposure to maternal common mental illness and maternal infection in the pregnancy cohort. Maternal and child characteristics were similar for cohorts exposed at 1-and 2-years. Maternal common mental illness and maternal infection in pregnancy were both associated with lower maternal age, maternal smoking, low birthweight, elective and emergency Caesarean section. Maternal mental illness was associated with infant resuscitation at birth. Women with depression or anxiety were more likely to experience infection during pregnancy or at any other exposure period.

**Table 1.**
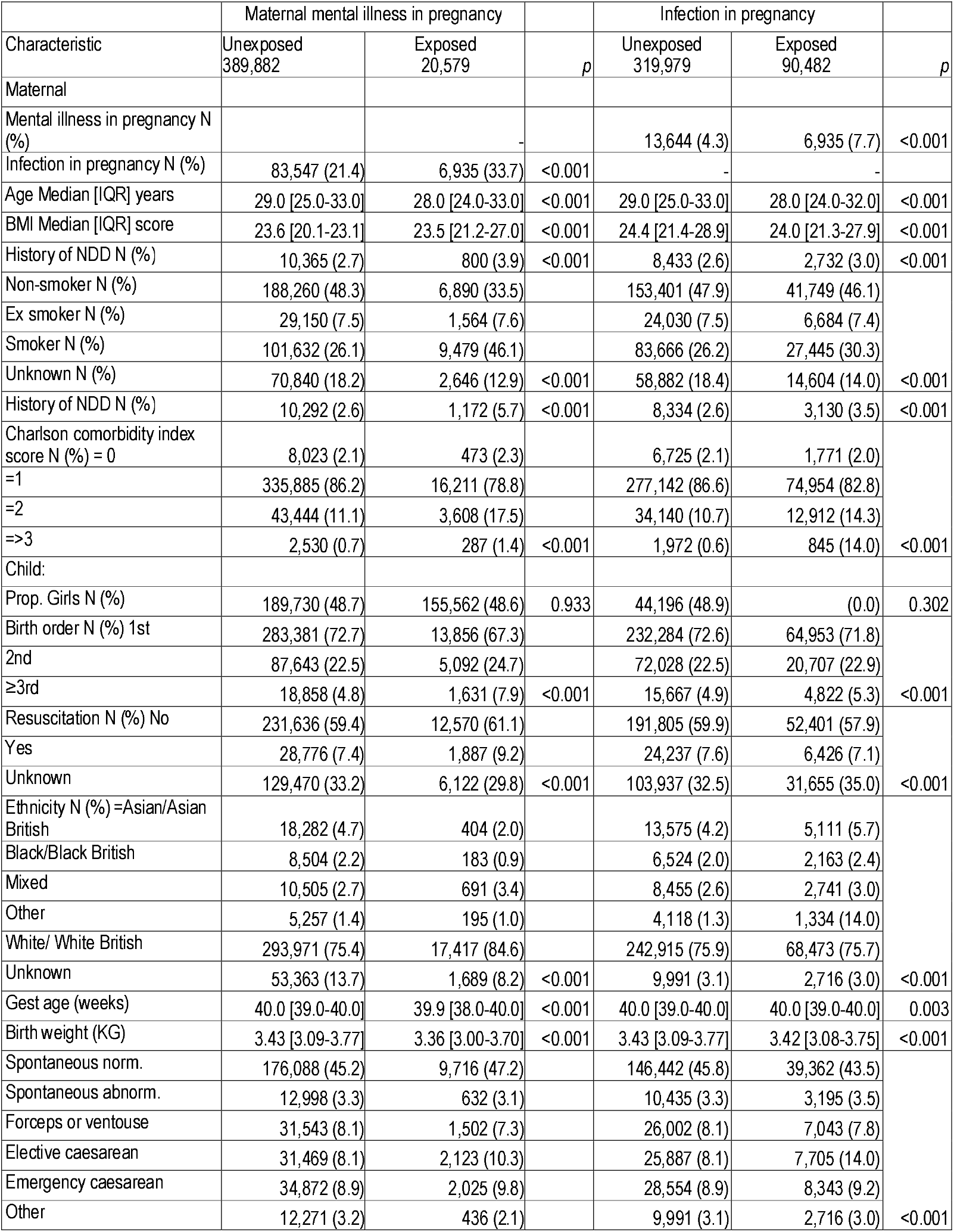
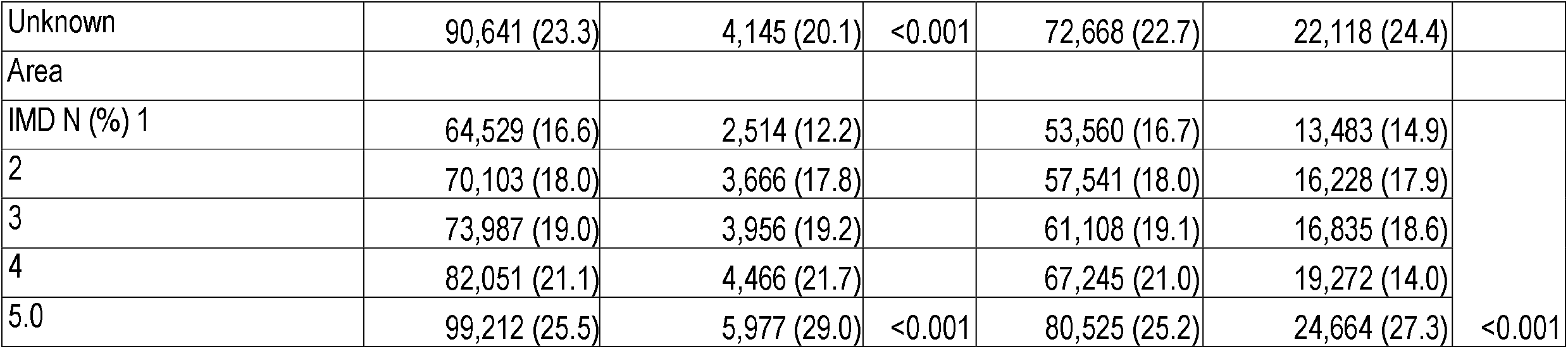
The characteristics of women and their children unexposed and exposed to mental illness and infection in pregnancy.

Association between maternal mental illness or maternal infection exposure in pregnancy and offspring NDD compared to exposure at other times.

Maternal mental illness during pregnancy increased offspring NDD risk (5.53 in exposed versus 3.09 events per 1000 person-years in unexposed offspring, HR (Hazard Ratio) = 1.83, 95%CI 1.69-1.99); the hazard ratio attenuated after adjustment for maternal age, maternal smoking, comorbidities, BMI, history of maternal NDD and offspring gender, ethnicity, birth year, region, and level of deprivation (adj HR=1.58, 95%CI 1.46-1.72). The unadjusted and adjusted association of exposure to maternal mental illness one year (5.02 versus 3.01 events, HR=1.70, 95%CI 1.59-1.81, adj HR=1.49, 95%CI 1.39-1.60) and two years prior to pregnancy (5.34 versus 3.03 events, HR=1.79, 95%CI 1.67-1.93, adj HR=1.62, 95%CI 1.50-1.74) were like risk following exposure during pregnancy. Compared to children unexposed to maternal infection in pregnancy, exposed children had an increased risk of offspring NDDs, but to a lesser extent than exposure to maternal mental illness during pregnancy (3.68 versus 3.06 events per 1000 person-years, HR=1.21, 95%CI 1.15-1.27). After adjusting for all potential confounders, this effect was reduced (adj HR= 1.16, 95%CI 1.10-1.22). There was also an association between maternal infection and offspring NDDs following maternal infection one year (3.74 versus 3.01, HR=1.25, 95%CI 1.19-1.32, adj HR=1.20, 95%CI 1.14-1.27) and two years prior to the index pregnancy (3.75 versus 3.05, HR=1.24, 95%CI 1.17-1.31, adj HR=1.19 95%CI 1.12-1.25).

The association between maternal mental illness and offspring NDD was retained after adjustment for confounders and pregnancy exposure to antidepressant (adj. HR=1.21, 95%CI=1.07-1.37), as was the association between infection in pregnancy and NDD after adjustment for antibiotic use (adj. HR=1.07, 95%CI 1.02-1.12).

Similar patterns of associations were observed for each type of NDD outcome (see Figure 1) and did not vary by gender of child (see eFig 2). Effects did not vary by trimester (see Figure 2).

**Figure 1.**
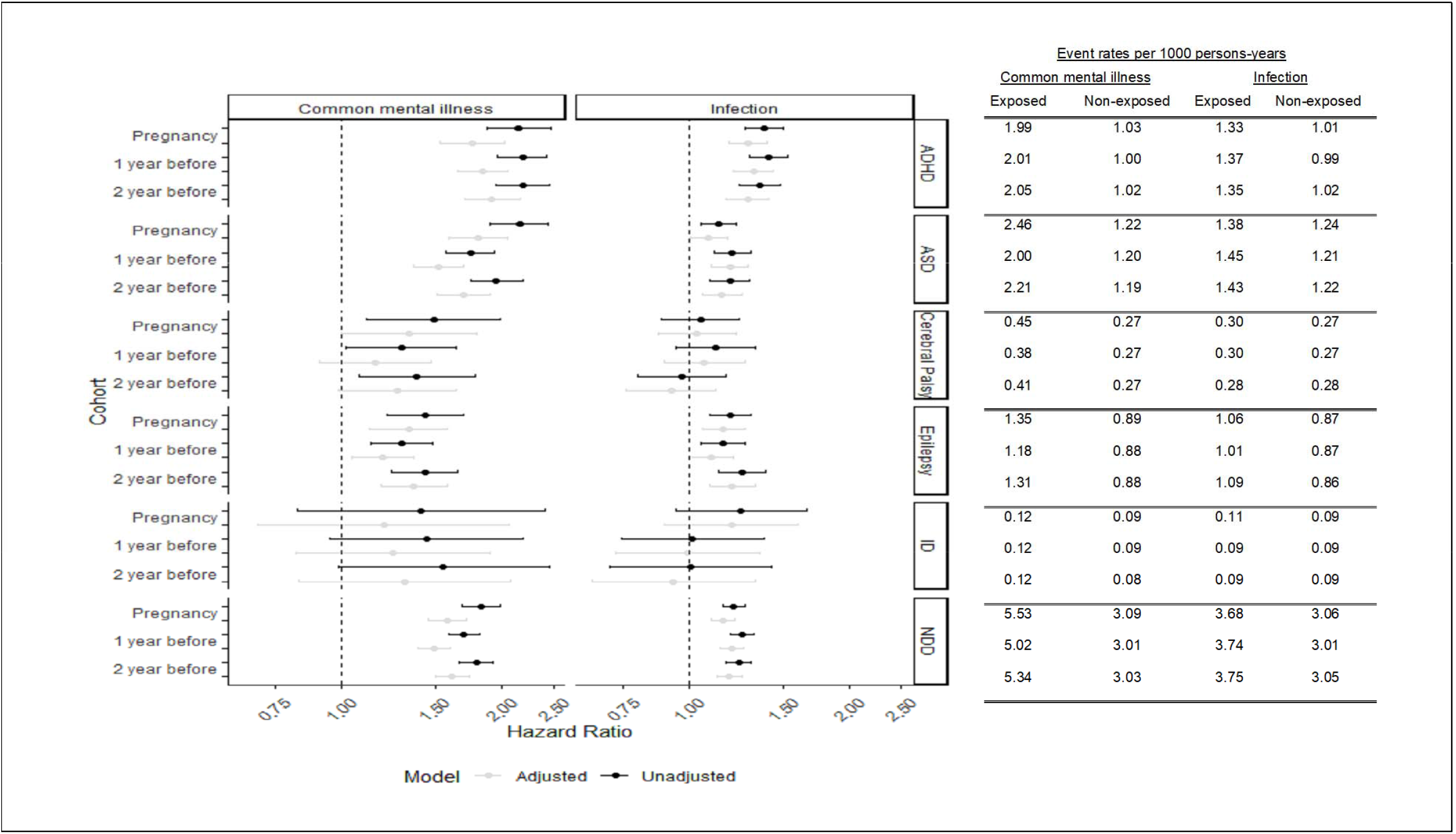
The association between maternal mental illness or infection and NDD outcome for pregnancy, 1 year prior and 2-year prior cohorts. Adjusted model includes: maternal; age, smoking status, comorbidities, BMI, history of NDD; child; gender, ethnicity, birth year; and area; region and level of deprivation, maternal mental illness adjusted for maternal infection and maternal infection adjusted for maternal mental illness.

**Figure 2.**
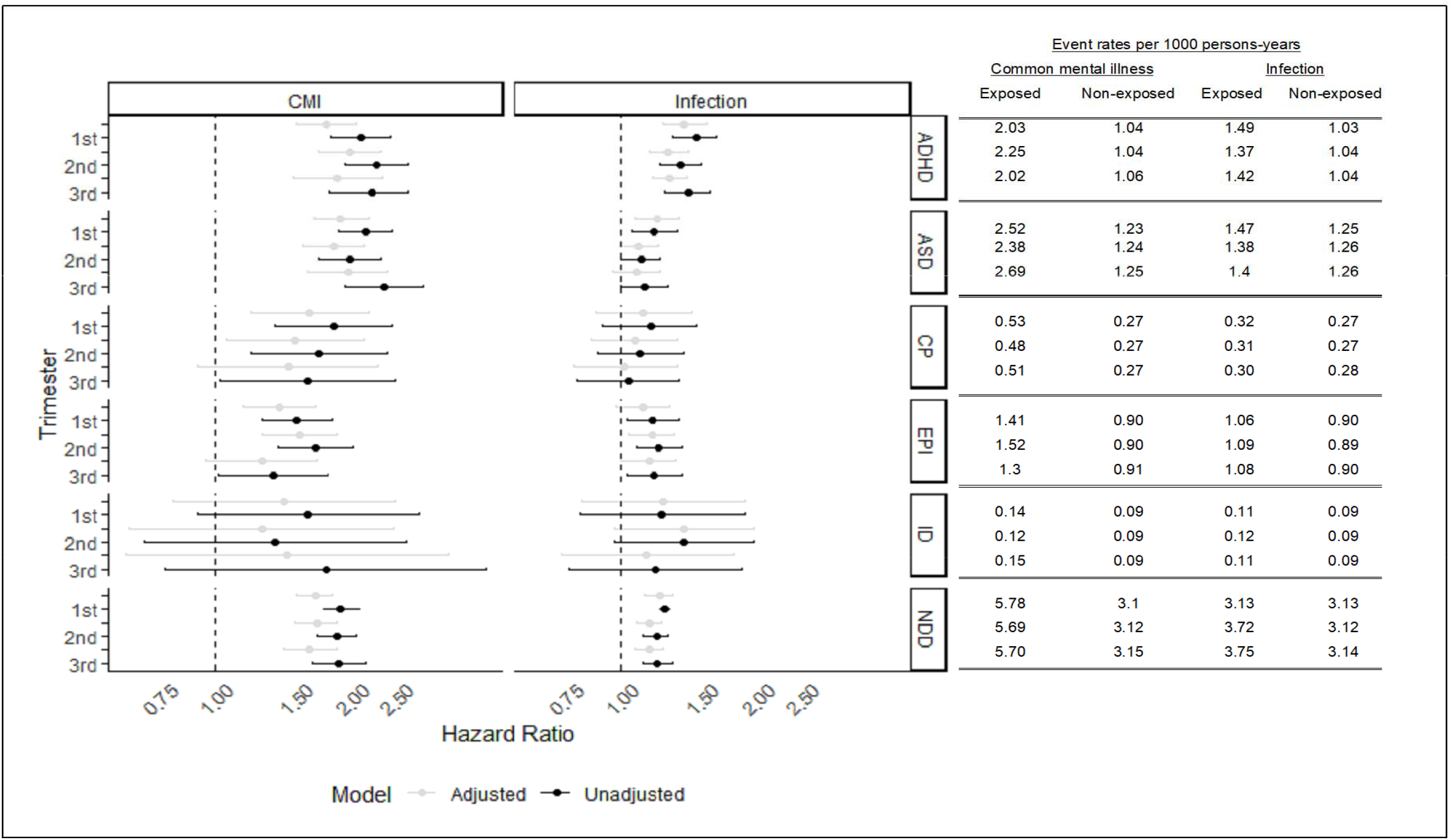
The effect of mental illness and infection in pregnancy and NDD, by pregnancy trimester. Model adjusted for; maternal; age, smoking status, comorbidities, BMI, history of NDD; child; gender, ethnicity, birth year; and area; region and level of deprivation, maternal mental illness adjusted for maternal infection and maternal infection adjusted for maternal mental illness.

### Discordant sibling analysis

We examined 7,738 siblings discordant for exposure to maternal mental illness and 60,534 siblings discordant for exposure to maternal infection during pregnancy. Rates of NDD were similar for exposed and unexposed siblings (5.66 versus 5.36 events per 1000 person years). After adjustment for maternal age, smoking status, BMI, alcohol, or substance misuse, Charlson comorbidity index, and offspring gender, ethnicity, birth year and birth order, there was no increase associated with maternal mental illness (adj HR=0.97, 95%CI 0.77-1.21); a similar pattern was observed for offspring ASD (adj HR=1.01, 95%CI 0.71-1.43) and ADHD (adj HR=0.92, 95%CI 0.65-1.31). There was no evidence of a difference in rates of NDD among siblings exposed and unexposed to maternal infection during pregnancy (3.44 versus 3.44, adj HR=0.99, 95%CI 0.90-1.08). This pattern of results was repeated for ADHD (adj HR=1.05, 95%CI 0.89-1.23) and ASD (adj HR=0.99, 95%CI 0.86-1.15).

Results were similar whether siblings were sisters or brothers (see Figure 3).

**Figure 3.**
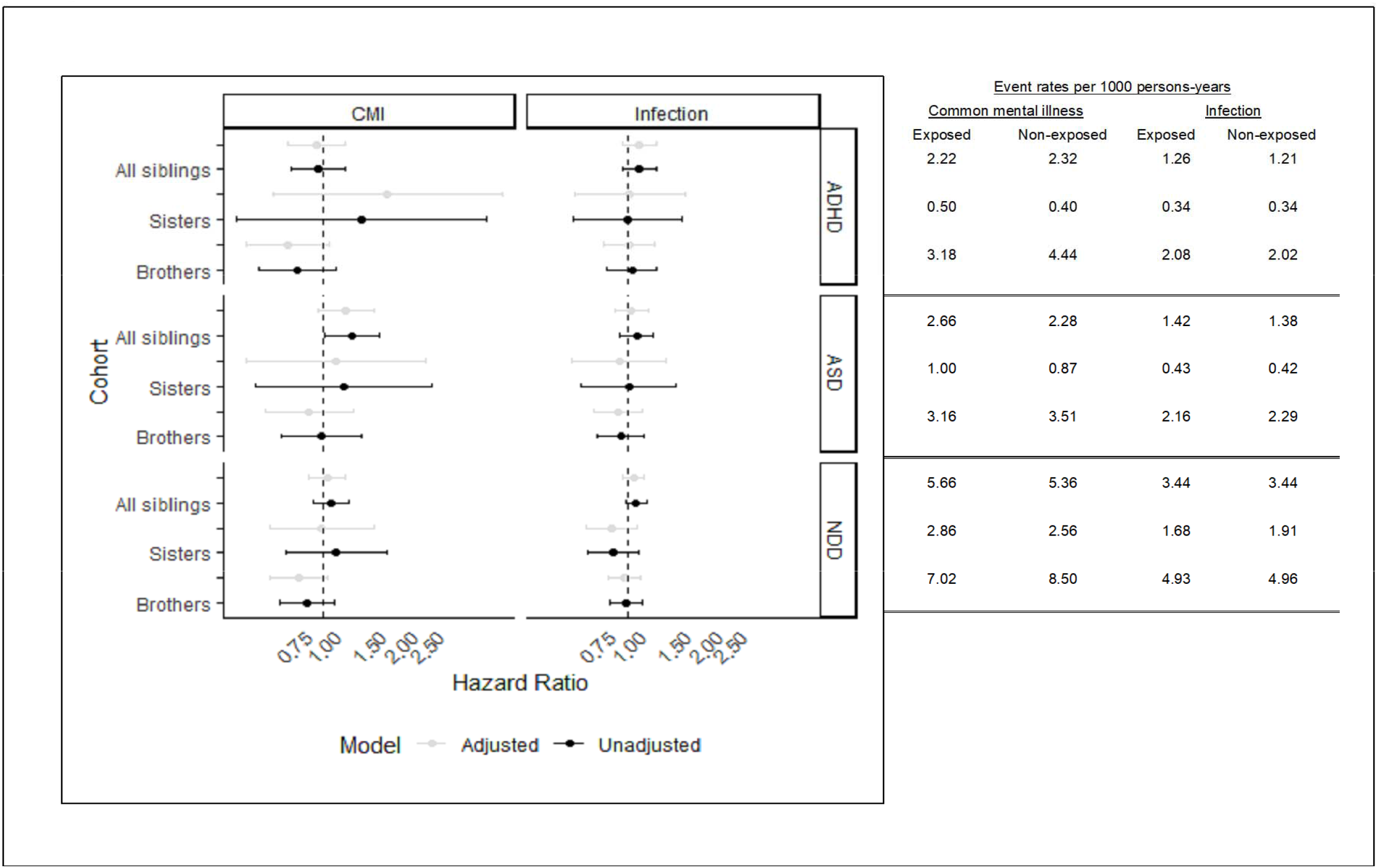
Sibling association for autism/ASD and ADHD, for all siblings, sisters and brothers. Model adjusted for; maternal; age, smoking status, comorbidities, BMI; child; gender, ethnicity, birth year and order.

### Sensitivity analyses

The association between maternal mental illness (HR=1.83, 95%CI 1.69-1.99) and maternal infection (HR=1.21, 95%CI 1.15-1.27) in pregnancy and NDD remained highly elevated for secondary care diagnoses only. When we excluded children with delayed registration at their GP practice (which may select families with more dysfunction), the association between maternal mental illness and maternal infection in pregnancy and offspring NDD did not change. Nor did these exclusions alter the pattern of attenuation of effects after each adjustment (confounders, exposure in other periods).

Modelling maternal mental illness exposure as episodic increased the number of women with mental illness during pregnancy to 36,439 (8.8%). This did not materially change the results.

## Discussion

We constructed a ‘natural experiment’ using high quality, population-based data from primary and secondary care sources to create different exposure windows for examining the relationship between exposure to maternal mental illness or maternal infection and offspring neurodevelopment. Overall, a child whose mother experienced mental illness or infection during pregnancy had higher incidence of NDD in childhood. However, this increase was similar whether exposure occurred during, or 1-2 years before onset, of pregnancy. Nor did risk vary by trimester, counter to MiA hypothesised mechanism of action. We report similar risks for siblings whose mothers had neither recorded mental illness nor infection during pregnancy. Taken together, these findings strongly imply that there is no specific link between maternal mental illness or maternal infection during pregnancy and subsequent risk of offspring NDD.

### Research in context

The observational literature examining associations between antenatal (severe) maternal stress, offspring brain development and/or cognitive delay is inconsistent (Abel et al., 2014; Khashan et al., 2008). Timing of offspring exposure does not alter the association between exposure and NDD risk in children, which places doubt on the proposed mechanism of risk transmission triggered in pregnancy (Arcangeli et al., 2012; Brynge et al., 2022; O’Connor & Ciesla, 2022). Moreover, sibling analyses and our findings that maternal infection outside of pregnancy also increased offspring NDD risk point to shared familial susceptibility to common mental illness, infection and NDD within families consistent with previous reports (Han et al., 2021; Lee et al., 2015; Lydholm et al., 2019; Qiao et al., 2020). The sibling analyses account for half of the unmeasured genetic effects, and potentially a greater proportion of shared environment if the children are born close together. The marked regression to the mean suggests factors with known links to lower IQ such as family deprivation/poverty and intimate partner violence must be considered as candidates for environmental confounding (Abel et al. 2019b). Larger genome wide association studies also fail to support the MiA hypothesis: genes associated with susceptibility to infection are elevated in adults and children with neuropsychiatric and neurodevelopmental disorder (Karlsson, Sjöqvist, Brynge, Gardner, & Dalman, 2022; Nudel et al., 2019).

### Strengths and Limitations

Several strengths of this study help extend the current literature. First, this large population sample captures exposure to maternal mental illness and infection in the community and not at hospital admission, meaning it represents a more accurate exposure estimate than prior analyses (Brynge et al., 2022). Furthermore, women might be admitted to hospital because of medical concerns about the pregnancy (which could independently influence NDD risk) and may develop an infection in hospital (Lee et al., 2015). Second, the large sample size and statistical power means we were able to capture the incidence of a variety of neurodevelopmental disorders and examine effects by trimester. In the UK, considerable efforts have been made to reduce the stigma of reporting mental health problems, and in the perinatal period women are asked about their mental health when they attend antenatal appointments, meaning incidence in pregnancy should be well recorded. In a prior analysis using a similar cohort of mothers we report higher rates of mental illness in the UK (Abel et al., 2019b) than Sweden (Pierce et al., 2020), where rates were extracted from primary and secondary care datasets, suggesting primary care captures more of the mental health risk in communities. Thirdly, triangulation, where the findings of two studies whose methods are independent are combined. is endorsed specifically in its application to causal inference (Ohlsson & Kendler, 2020).

Despite this, reliable identification of illness in healthcare records can be challenging (Abel et al., 2019b), particularly identifying the exact date of diagnosis/illness onset. This means that some women and offspring might be misclassified as unexposed particularly those with milder disorder. We think our slight underestimate of exposure outcome incidence will not have contributed to an exaggerated association. We adjusted the measurement of depression, anxiety and NDD to take account of delayed presentation to services and this did not change our results. Finally, maternal infection in pregnancy is common. In our sample, 90,482 (22%) of pregnant women presented to primary care with an infection, higher than other studies of infection. That said, many infections in pregnant women are mild and self-limiting and do not require treatment, meaning our primary care contacts are likely to underestimate the true incidence of maternal infection because many women will not seek help, or remain unaware they have an infection.

### Future Research

Whilst our finding that developing a common mental illness or infection in pregnancy severe enough to require treatment does not cause NDD in your offspring is informative to people planning or who are pregnant and those who work with them, future innovative study designs that employ negative controls including using serology, might further inform causality for specific infections. For example, in a prior analysis, we reported that people who tested negative for COVID-19 infection were at similar risk of mental illness during the pandemic as those who tested positive (Abel et al., 2021).

Whilst electronic health records are useful for assembling large cohorts, they may miss milder cases, where the family context might affect the timing of diagnosis. For clinical and public health reasons, it is vital to understand whether maternal infection materially contributes to risk of atypical offspring neurodevelopmental (Ji-Xu & Vincent, 2020). Comparing the mean difference in validated scales of developmental delay might be more robust, but currently these types of data are not readily available in datasets of the size required to advance our understanding in this area. In a much smaller sample (Shuffrey et al., 2022) demonstrated that pregnancy exposure to the COVID-19 pandemic, but not to COVID infection per se was associated with offspring developmental delay assessed through mean scores from the All-Stages’ Questionnaire.

## Conclusions

Our findings should reassure mothers with mental illness – especially given their excess likelihood of infection during pregnancy-as well as mothers of children with NDD who may have considered themselves to ‘blame’ because of pregnancy exposures. Our findings clearly indicate that timing of maternal mental illness or infection is not associated with offspring neurodevelopmental risk.

Where it is unethical or unfeasible to manipulate exposures, using population datasets may help refine the implications of more traditional observational findings or, indeed, of Fmri studies of prenatal stress which often cannot account for confounding, or are significantly biased by small sample sizes (Wu et al., 2022).

## Supporting information

etable

## Data Availability

All data are available only once approval has been obtained through the individual constituent entities controlling access to the data. The primary care data can be requested via application to the Clinical Practice Research Datalink, secondary care data can be requested via application to the hospital episode statistics from the UK Health and Social Care Information Centre.

https://github.com/HollyHope/NDD_CPRD

## Acknowledgements

This study is based in part on data from the Clinical Practice Research Datalink (CPRD) obtained under licence from the UK Medicines and Healthcare products Regulatory Agency. The study was approved by the Independent Scientific Advisory Committee (ISAC) for MHRA (Medicines and Healthcare products Regulatory Agency) Database Research (protocol number: 17_187). Generic ethical approval for observational research using CPRD with approval from ISAC has been granted by a Health Research Authority (HRA) Research Ethics Committee (East Midlands—Derby, REC reference number 05/MRE04/87).

The data is provided by patients and collected by the NHS as part of their care and support. Hospital Episode Data Copyright © (2017), are re-used with the permission of The Health & Social Care Information Centre. All rights reserved.

## Data sharing

Data sharing: Read codes used are published on Clinicalcodes.org. Electronic health records are, by definition, considered “sensitive” data in the UK by the Data Protection Act and cannot be shared via public deposition because of information governance restriction in place to protect patient confidentiality. Access to data are available only once approval has been obtained through the individual constituent entities controlling access to the data. The primary care data can be requested via application to the Clinical Practice Research Datalink (www.cprd.com/researcher), secondary care data can be requested via application to the hospital episode statistics from the UK Health and Social Care Information Centre (www.hscic.gov.uk/hesdata).

## Ethical Standards

The authors assert that all procedures contributing to this work comply with the ethical standards of the relevant national and institutional committees on human experimentation and with the Helsinki Declaration of 1975, as revised in 2008.

## Competing interests

The authors declare none.

## Notes

### Competing Interest Statement

The authors have declared no competing interest.

### Funding Statement

This study was funded by the European Research Council (ERC).

### Author Declarations

The Independent Scientific Advisory Committee (ISAC) of the Medicines and Healthcare products Regulatory Agency gave ethical approval for this work.

