## Supplementary material for "The causal association between maternal depression, anxiety and infection in pregnancy and neurodevelopmental disorders among 410,461 children- a population study using quasi-negative control cohorts and sibling analysis": etable

Supplementary tables and figures

1.) eFig1: Consort figure
2.) eTable1: Comparison of risk cohorts
3.) eTable2: Description of outcome NDD codes by data source
4.) Exposure algorithm
5.) eFig2: The association between maternal exposure to mental illness or infection and autism/ASD, ADHD, cerebral palsy, intellectual disability and epilepsy for boys and girls

Singleton births registered at a HES linked CPRD practice 01.01.90-31.12.17 715,421

Excluded =169, 012 Good quality data collection began after child's DOB

Excluded =1928 Mother transferred out of the practice before the child DOB

Excluded =134,110 No delivery record in the mother's HES

Pregnancy cohort= 410,694 Mother in record 280 days prior to DOB

1 year prior = 352,993, Mother in record 1 year +280 days prior to DOB

2 years prior= 303,016 Mother in child's record 2 years + 280 days prior to DOB

eFig1: Consort figure

eTable1: Characteristics of cohorts

|  | | **Pregnancy** | **1 year before** | **2 years before** |
| --- | --- | --- | --- | --- |
| **Common mental illness N (%)** | Exposed | 20,579 (5.0) | 34,438 (9.8) | 27,840 (9.2) |
| **Infection N (%)** | Exposed | 90,482(22.0) | 82,062 (23.3) | 70,011 (23.2) |
| **Maternal age at start** | Median [IQR] | 30 [26-34] | 29 [25-33] | 28 [24-32] |
| **Maternal BMI** | Median [IQR] | 24.1 [21.5-28.0] | 23.7 [21.2-27.3] | 23.6 [21.1-27.1] |
| **Charlson comorbidity index N (%)** | 0 | 8,773(2.1) | 7,584 (2.2) | 5,892 (2.0) |
|  | 1 | 348,321 (84.9) | 299,433 (85.2) | 259,175 (85.9) |
|  | 2 | 50,289(12.3) | 42,034 (12.0) | 34,741 (11.5) |
|  | 3 or more | 3,078(0.8) | 2,563 (0.7) | 1,971 (0.7) |
| **Smoking status N (%)** | Never smoked | 199,973(48.7) | 171,132 (48.7) | 143,229 (47.5) |
|  | Ex-smoker | 33,223(8.1) | 26,056 (7.4) | 20,060 (6.7) |
|  | Current smoker | 116,295 (28.3) | 100,431 (28.6) | 82,942 (27.5) |
|  | Not known | 60,970 (14.9) | 53,995 (15.4) | 55,548 (18.4) |
| **Mother NDD N (%)** | Any | 11,464 (2.8) | 9,758 (2.8) | 8,449 (2.8) |
| **Child gender N (%)** | Female | 199,758 (48.7) | 171,200 (48.7) | 146,908 (48.7) |
| **Ethnicity N (%)** | Asian/British Asian | 18,686 (4.6) | 14,356 (4.1) | 11,260 (3.7) |
|  | Black/ black British | 8,687 (2.1) | 6,733 (1.9) | 5,229 (1.7) |
|  | Mixed | 11,196 (2.7) | 9,260 (2.6) | 7,766 (2.6) |
|  | Other | 5,452 (1.3) | 4,104 (1.2) | 3,170 (1.1) |
|  | White | 311,388 (75.9) | 270,748 (77.0) | 235,480 (78.0) |
|  | Unknown | 55,052 (13.4) | 46,413 (13.2) | 38,874 (12.9) |
| **IMD N (%)** | 1 | 67,043(16.3) | 56,338 (16.0) | 46,942 (15.6) |
|  | 2 | 73,769 (18.0) | 62,795 (17.9) | 53,338 (17.7) |
|  | 3 | 77,943 (19.0) | 66,462 (18.9) | 56,754 (18.8) |
|  | 4 | 86,517 (21.1) | 74,341 (21.1) | 64,145 (21.3) |
|  | 5 | 105,189 (25.6) | 91,678 (26.1) | 80,600 (26.7) |
| **Child NDD N (%)** | Any | 11,165 (2.7) | 9,637 (2.7) | 8,426 (2.8) |
| **Gestational age** | Median [IQR] | 40 [39-40] | 40 [39-40] | 40 [39-40] |
| **Birthweight N (%)** | <2000g | 4,887 (1.2) | 4,234 (1.2) | 3,684 (1.2) |

eTable2: Description of outcome NDD codes by data source

|  | **Data source** | | |
| --- | --- | --- | --- |
| **Outcome** | **CPRD prescriptions** | **HES diagnosis at Outpatient visit** | **HES ICD10 diagnosis at hospital admission** |
| **ADHD** | BNF chapter 4, section 4, paragraph 0 (excluding caffeine) | F84 | F84 |
| **ASD** | NA | F90 | F90 |
| **ID** | NA | F70-F79 | F70-F79 |
| **Cerebral Palsy** | NA | G40 | G40 |
| **Epilepsy** | NA | G80-G83 | G80-G83 |

The CPRD read codes for ADHD, ASD, ID, CP and epilepsy are available at github.

Mental illness exposure algorithm

The algorithm considers common mental illness to be episodic, and takes account of the reality that a diagnosis of depression or anxiety will only be entered once into a person’s primary care record. Therefore mental illness within a given period (in pregnancy, 1 year or 2 years before pregnancy) was calculated using the following rules:

1. Record of a diagnosis of anxiety or depression in the period.
2. Record of a prescription to treat anxiety or depression in the period, where there was a historic diagnosis of anxiety or depression.
3. Record of a symptom of anxiety or depression in the period, where there was a historic diagnosis of anxiety or depression.
4. Record of a symptom of anxiety or depression within 3 months of a prescription for anxiety or depression in the period.

See [github](https://github.com/HollyHope/NDD_CPRD) for list of exposure prescriptions, symptoms, and diagnoses.

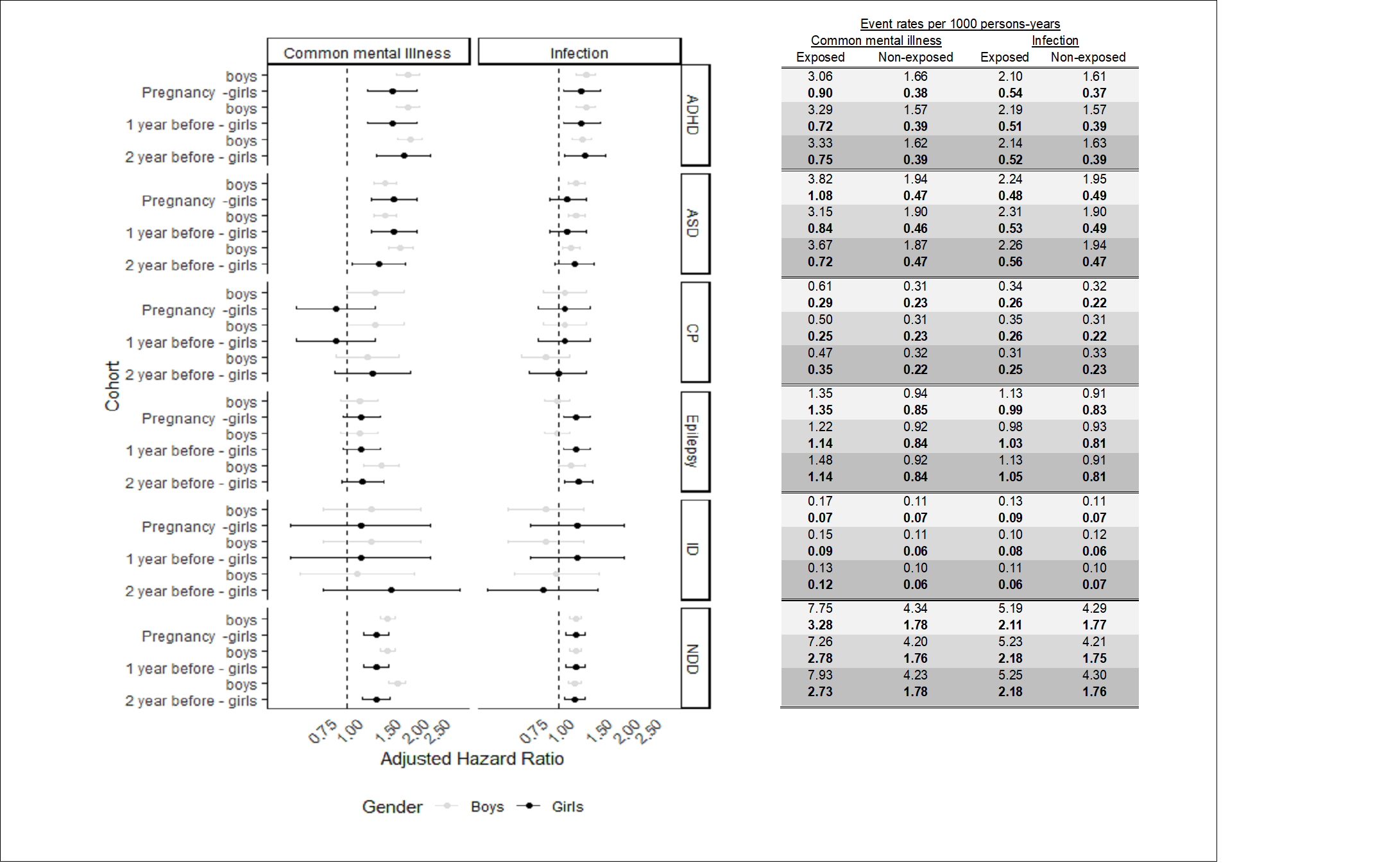

eFig2: The association between maternal exposure to mental illness or infection and autism/ASD, ADHD, Cerebral palsy, Intellectual disability and Epilepsy for boys and girls

Model adjusted for; maternal; age, smoking status, comorbidities, BMI, history of NDD; child; gender, ethnicity, birth year; and area; region and level of deprivation. Maternal mental illness adjusted for maternal infection and maternal infection adjusted for maternal mental illness.
